# Can Chinese Herbal Medicine Be Useful in Treatment of Type 2 Diabetes Mellitus? A Systematic Review of Medicinals Used and Clinical Impact

**DOI:** 10.1101/2025.02.18.25322375

**Authors:** Anisa Finney, Lisa Conboy

## Abstract

Purpose: With Diabetes Mellitus (DM) on the rise, biomedicine has identified its treatment as challenging due to the multitude of interventions necessary for disease management^1^. Biomedical treatment utilizes a single action approach, placing patients in the care of one specialist at a time. The traditional Chinese Herbal Medical approach can be of great utility to DM patients administered by a single practitioner aiming to address whole body imbalances. This project synthesized available research discussing Chinese Herbal medicine in the treatment of T2DM via PubMed. Methods: Systematic review was used to search PubMed database October 2022-April 2023 leading to eventual retrieved sample of 30 articles. Search terms included *Chinese Medicine* AND *Diabetes Mellitus* OR *Chinese Medicine* AND *Type 2 Diabetes* with the filters of clinical trial, open access, in English. Results: Thirty articles met the inclusion criteria and were included in this review. Chinese herbal medicines have been extensively researched appearing in multiple forms such as pills, single herbs, granules, and decoctions. Of the 30 studies, the most reported outcomes were reduced fasting blood glucose, reduced HbA1c, and improved glycemic control. Conclusions: Further research is necessary to capture the role of Chinese herbal medicine in the integrative treatment of T2DM patients.

## Introduction

Diabetes Mellitus is a metabolic disease most easily distinguished by poor control of glucose levels in the bloodstream^1^. While there are multiple subcategories, Diabetes Mellitus Type 1 and Type 2 are the major differentiations. According to the National Center for Health Statistics, Diabetes Mellitus is the eighth leading cause of death in the United States^2^, and the ninth leading cause of death in the world^3^. Diabetes Mellitus Type 1 is generally characterized by insulin absence and Type 2 by insulin resistance, both precede hyperglycemia. Hyperglycemia, or high blood sugar, is characterized by high levels of glucose in the blood, typically 125 to 130 mg/dL^4^. Hyperglycemia occurs where the body lacks sufficient insulin or cannot properly use it to transport glucose into the blood. Chronic hyperglycemia leads to glycation of lipids and proteins manifesting as damage to the kidneys, peripheral nerves, and collateral blood vessels in the retina. Damage caused by glycation eventually leads to classic Diabetes Mellitus complications such as neuropathy, retinopathy, nephropathy, diabetes-related blindness, dialysis, and limb amputation^1^. The objective of this literature review is to become familiar with the breadth of research available on this topic and analyze findings to guide future research to better understand the research question (Can Chinese Herbal Medicine Be Useful in Treatment of Diabetes Mellitus) and assist medical communities in establishing the most effective plan of treatment for patients with this disease.

### Biomedical Treatment for Type 2 Diabetes Mellitus

The standard of care treatments for T2DM (see Table 1) in biomedicine are 1) educating healthy eating habits, 2) regular exercise and weight loss, 3) blood sugar monitoring and diabetes medication, and 4) insulin therapy^5^. Healthy eating habits refers to reducing intake of refined carbohydrates, starchy vegetables and refined sugars and focusing on high fiber, whole foods. Exercise recommendations include aiming for 150 minutes or aerobic exercise and two to three resistance training sessions per week (adults). Weight loss is emphasized because lower BMI is associated with greater blood glucose stability. Commonly used medications are antidiabetic medications to control blood sugar such as Metformin, statins to address hyperlipidemia, and when the former medications are adequate, insulin injections are used^5^. Blood sugar monitoring includes educating the patient how to use a finger stick glucose monitor or a continuous glucose monitor to periodically track their glucose levels (i.e., before and after meals, in the morning and before bed) and help better tailor treatment plans. New medications are often implemented with T2DM patients, but those mentioned above are typically used first.

**TABLE 1:**
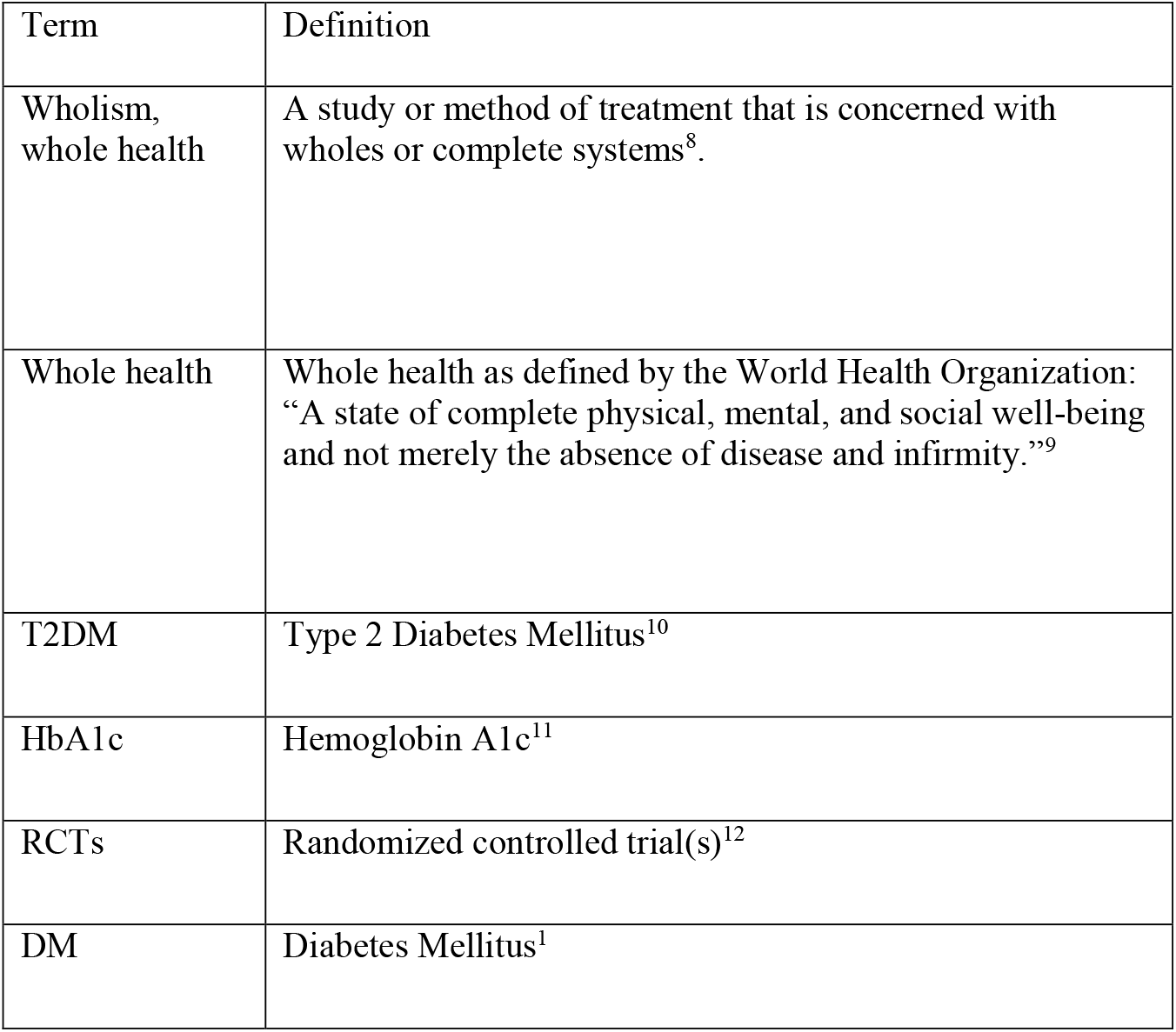
Terms.

## Background

This condition is worthy of discussion because biomedical models of treatment often involve a single action approach, placing patients in the care of one specialist at a time to work on a single physiological system, such as an endocrinologist to address hormone-related conditions or a nephrologist to address kidney functioning. The biomedical community has identified treatment of Type 2 Diabetes Mellitus as challenging due to the necessity of multiple interventions to manage disease symptoms. By parsing treatment between various specialists with no central line of communication, long term healing is often more difficult to attain. EAM medicine is a wholistic system and may offer better integration of care for the whole-body system.

The term in Chinese best characterizing Diabetes Mellitus is “xiaoke”. Xiaoke signifies a type of metabolic disease with a multitude of interwoven pathological mechanisms underlying the condition^6^.

Traditional Chinese Medicine treatment of Diabetes Mellitus involves a multi-faceted and whole-body approach. Traditional Chinese Medicine has been rooted in the whole-body approach for centuries and may be a suitable fit to combat a global disease affecting countless individuals. Formulas used in Chinese Herbal medicine include a multitude of herbs, carefully selected according to actions in the body, as well as how these herbs will interact with one another. Herbs can be added to balance actions of another to achieve desired effects. The personalized nature of Chinese Herbal medicine means practitioners not only reach a deep understanding of the condition a patient is presenting with, but also their individual constitution. Chinese medicine does not have established ‘standard of care’ for a biomedical condition, rather each patient receives and individualized treatment. The synergistic effects achievable by this individualized medicine could be of great utility toward establishing a comprehensive treatment plan without parsing care among a team of different specialists.

## Methods

Using the PubMed search criteria of “Chinese Medicine” AND “Diabetes Mellitus” or “Chinese Medicine” AND “Type 2 Diabetes”, all with the filter of “clinical trial”. Studies were then filtered to include studies where Chinese herbal medicine was used to address symptoms of T2DM (such as hyperglycemia). This PubMed search was conducted between October 2022 and April 2023. Inclusion criteria search terms: clinical trial, randomized controlled trial, systematic or multicenter reviews, experimental group taking Chinese herbal medicine, control group (as comparison group not taking Chinese herbal medicine), biomedical diagnosis of T2DM or impaired glucose tolerance, tracking main diabetes markers before and after trial (such as blood glucose, hemoglobin A1c, 2-hour post-prandial fasting glucose, fasting blood glucose, etc.), and clearly stated results. Exclusion criteria: Article not available in English, no full-text access, focus on pharmacological actions of single herbs or herbal formulas, animal studies, focus on single symptoms or complications associated with T2DM. This review was formatted in accordance with the PRISMA guidelines for systematic reviews and acupuncture. The Lancet’s formatting guidelines for systematic reviews were also followed. Author AF performed the initial review and author LC acted as the second reader. The results of this search are summarized by the included flow chart in Figure 1.

**Figure 1.**
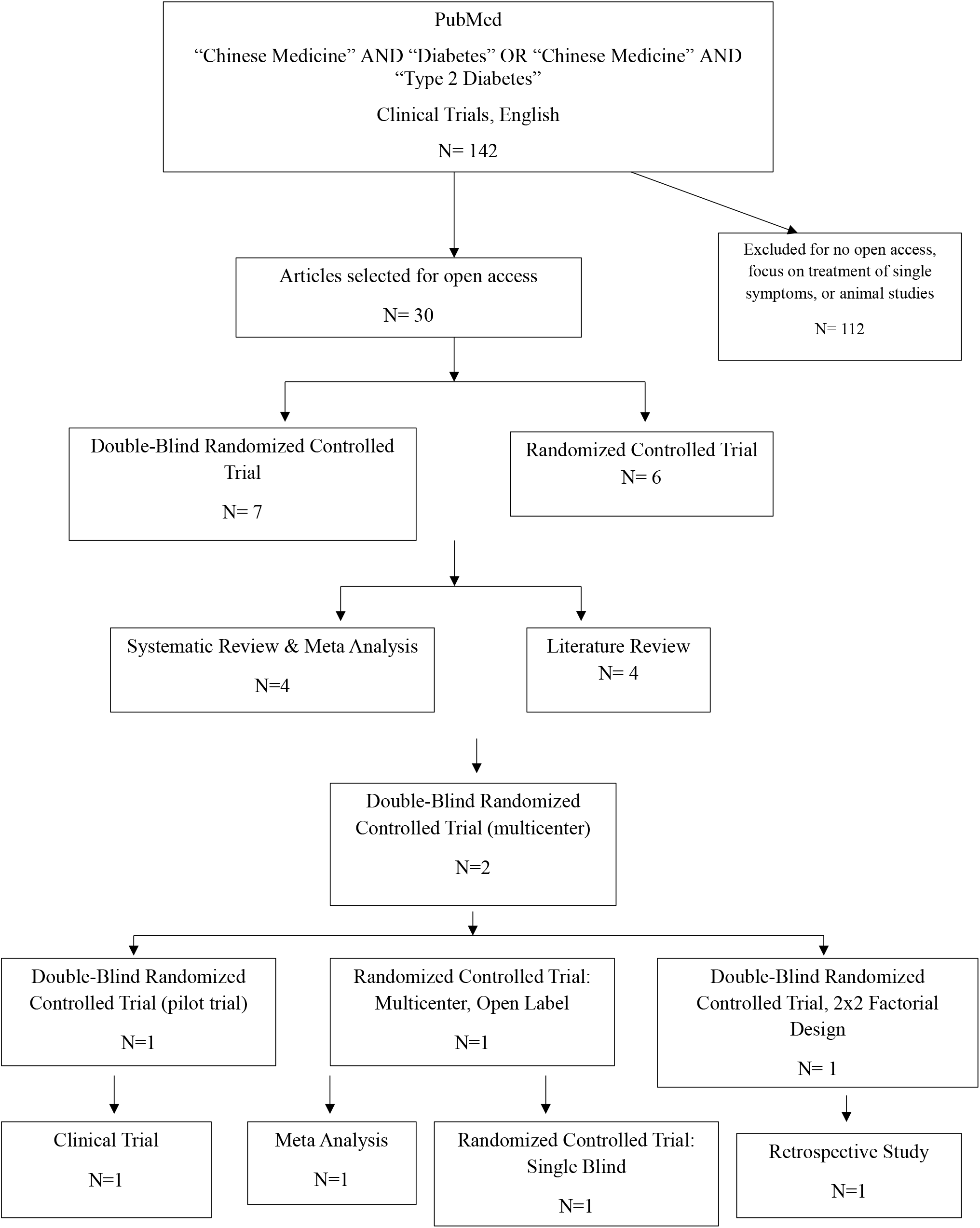

### Study Risk of Bias Assessment

This review utilized publications strictly from articles published in English, and those which were accessible to the public (did not require institutional affiliation or payment to access). use of only free articles in English may have prevented the authors from seeing the whole picture of the current state of research, and how Chinese Herbal medicine has been implemented to treat patients with Type 2 Diabetes Mellitus. Perhaps those restricted access studies propose more nuisance, or the language restriction may have lay hidden studies implement of Chinese Herbal medicine in a more classic or traditional way. Bias could occur in the form of believing there has been no prior research done using customized Chinese Herbal medicine and that creating a study on that topic would be entirely novel, when in fact, there may already be one, or multiple studies currently published on the topic in restricted access literature.

## Results

Thirty publications were identified to fit the inclusion criteria mentioned above. These articles were analyzed in the fashion of a literature review. Details can be found in PDF document included as Figure 2 below.

This review has incorporated information from a total of 30 articles in English that met the inclusion criteria and offer full-text access. The research methodologies employed in these studies ranged from Double-Blind Randomized Controlled Trials, Systematic Reviews with Meta-Analysis, Literature Reviews, and Retrospective studies. The most common formats of research were the eleven Double-Blind RCTs (see Table 1) and the eight RCTs. These had an average study duration of 22 and 15 weeks, and an average 200 and 165 participants, respectively. Chinese herbal medicines were extensively considered, appearing in multiple forms such as pills, single herbs (specifically ginseng), granules, and decoctions.

Custom patient pills include ‘xiaoke pill’, ‘tianqi’, and ‘jiangtang xiaozhi’. These studies spanned from 1995 to 2022, demonstrating a sustained interest in the examination and utility of the effects of Chinese herbal medicine for glycemic control in T2DM patients.

### Risk of Bias in Studies

After careful analysis of the studies included in the review, Chinese Herbal approaches implementing wholism are absent. Studies relied on implementation of a single herb, or a single predetermined herbal formula which all participants received. Chinese Herbal medicine was intended to be practiced in a highly customized manner, accounting not only the current condition of the patient, but their constitution as well as variation in symptoms among patients with the same biomedical diagnosis. Failing to design and report data on a study implementing customized Chinese Herbal medicine for Type 2 Diabetes patients is failing to accurately assess the medicines’ ability to help enhance treatment in conjunction with biomedicine.

One avenue through which studies report safety is through the tracking of adverse events. Of the 30 trials included in this paper, six commented on presence or absence of adverse events as results of the study. As safety is of utmost importance in trials which include human subjects, as largely regulated by the Belmont Report^7^, if adverse events were not reported in the results, it is assumed that none occurred.

Columns ‘K’ and ‘L’ in the systematic review table served to outline the studies with authors affiliated with an East Asian Medicine Institution, and the studies which mentioned credentialed Acupuncturists/ East Asian Medicine practitioners among the authors. Of the 30 studies, 14 of them included authors affiliated with an East Asian Medicine Institution. Of the 30 studies, none mentioned credentialed Acupuncturists/ East Asian Medicine practitioners among the authors. Of the studies affiliated with an East Asian Medicine Institution, the average and median number of authors affiliated were seven and a half. As it pertains to the origin of the study, of the 30 studies, 20 of them had origins in East Asia, five of the 30 had origins in the United States, and seven of the 30 had origin outside East Asia and the United States. Motivation behind incorporating a section delineating authors affiliated with an East Asian Medicine institution as well as authors who held credentials of an acupuncturist (L.Ac.) or East Asian Medicine Practitioners (EAMP) or Doctors of Oriental Medicine (DOM) was fueled by curiosity. Curiosity as to the breadth of countries conducting research on this topic. Curiosity as to the studies which had affiliation with an East Asian Medicine institution and or any acupuncturist or East Asian Medicine Practitioners among the authors. The intention was not to purport that East Asia is the best and only place this research should be conducted, but rather to highlight all regions that have worked, or are working to contribute research in this field. Additionally, these columns serve to convey a wide-spread interest in the topic [Chinese herbal medicine as treatment for T2DM] across professions outside of East Asian Medicine. The final columns in the systematic review serve to demonstrate that quality and meaningful research can, and have, come from a range of countries.

### Significance

This review found numerous publications exploring effects of single Chinese medicinals or formulas and their utility in addressing common symptoms of complications of T2DM. Historical and systematic reviews of clinical trials were also included to understand the current breadth of research pertaining to treatment of T2DM patients through Chinese herbal medicine. Following initial review, treatment of the condition, past and present will be used to compile important findings to better equip current Chinese Medicine practitioners for treatment of an ever-increasing population of T2DM patients, especially in the west. This review intends to clearly define the role of Chinese Herbal medicine has played in treatment of this condition and help support the claim that Chinese medicine can support biomedical treatment to better manage patients with this disease.

### Discussion, Next Steps

The results of these studies provide valuable insights into the utility of Chinese herbal medicines in assisting the management of symptoms in T2DM sufferers. Across the 30 studies, the most common outcomes reported were reduced fasting blood glucose (FBG), reduced HbA1c, and improved glycemic control. Other outcomes included improved [2 hour] postprandial glucose, improved HDL cholesterol, enhanced hypoglycemic action of metformin and improved beta cell function. Despite these positive outcomes reported above, it is crucial to emphasize that further research is necessary to fully establish the role that Chinese herbal medicine can assume in treatment of T2DM. It is also necessary to use the literature to adequately address promising outcomes, it is crucial to note that further research is necessary to firmly establish the role Chinese herbal medicines can play in treating T2DM. It is also imperative to address the safety and potential side effects of these medicinals to garner higher acceptance in the biomedical community. Particularly, more investigation is needed on their potential as adjunctive care alongside conventional biomedical care for T2DM patients. The evidence thus far indicates potential benefits, but robust validation through future studies is necessary. An important limitation of these studies lies in the lack of wholism^8^ emphasized in treatment interventions. Although many studies reported favorable outcomes in T2DM patients treated with Chinese herbal medicine (single herb or a prepared patent), the thread of whole-system care distinct in East Asian Medicine was neglected. One cannot truly outline the utility of East Asian Medicine in this population of patients if other modalities are not also utilized. Some of these modalities include acupuncture, tui na body work, cupping, gua sha, qi gong practice as well as diet and lifestyle advice rooted in principles of East Asian Medicine. Of the bulk of research available about Chinese herbal medicine’s utility in treating Type 2 Diabetes Mellitus lies in the lack of studies implementing herbal medicine with customized formulas, fit to each patient’s constitution and presenting symptoms. Instead, studies rely on a standard of care treatment model, implementing the same intervention for all patients treated. To analyze EAMs true utility for T2DM patients, studies ought to be created and published which report on patients treated with customized formulas.

### Discussion on Credentials

Column ‘L’ in the systematic review, included to delineate studies which mentioned credentials Acupuncturists (L.Ac.) or East Asian Medicine Practitioners (EAMP), after careful analysis of the studies, was unremarkable. A few of the credentials mentioned in the studies were Doctors of Medicine (M.D.), Doctor of Philosophy (PhD) and one Masters of Medicine (MM), but no Acupuncturists, or East Asian Medicine Practitioners. But given that most studies did not display author credentials clearly in the publications, the presence or absence of these professionals cannot be assumed or denied at this time. In the future, it could add to the readers understanding, or to this field of research to highlight what type of professionals assisted in the study, especially as it pertains to discussion about whether all methods of

East Asian Medicine have been considered in treatment of T2DM patients. If licensed Acupuncturists or East Asian Medicine Practitioners have never been included among authors working in this field of study [Chinese Herbal treatment for T2DM, published in English, free article access], it could be advantageous in study development and analysis to have these professionals included in future publications.

This review cannot fully represent the state of publications on the given topic due to the exclusion of articles that were not open access (pay to access), or those published in a language besides English. To complete the review, search criteria must be expanded to explore publications which are pay to access and published in a language besides English. Failure to include publications from these sources could portray an inaccurate depiction of the current research using Chinese Herbal medicine in treatment for Type 2 Diabetes Mellitus.

## Discussion on Wholism

An important limitation of these studies lies in the lack of wholism emphasized in treatment interventions. Although many studies reported favorable outcomes in T2DM patients treated with Chinese herbal medicine (single herb or a prepared patent), the thread of whole-system care distinct in East Asian Medicine was neglected. One cannot truly outline the utility of East Asian Medicine in this population of patients if other modalities are not at least considered, if not utilized. Some of these modalities include acupuncture, tui na body work, cupping, gua sha, qi gong practice as well as diet and lifestyle advice rooted in principles of East Asian Medicine.

Future research should consider utilizing a cohort study model, which follows participants with similar characteristics (T2DM patients in this case) for a period. The study would center on Chinese herbal treatment through classic differential diagnosis, creating an individualized formula for each patient instead of a single herb or formula for all patients.none of the studies found as part of this review used this classic technique. The cohort study model could not only demonstrate the breadth of Chinese herbal applications, but to add to the biomedical model of one treatment for one condition and truly demonstrate the high specificity Chinese herbal medicine can contribute to treatment these patients. Such specificity is missing from current biomedical medical care and could be the missing component to constructing effective treatment plans to allow for better symptom relief and disease outcomes for T2DM patients. A use of tailored or individualized formulas, applied in a cohort research design, will both allow for greater customization of herbal formulas to best fit the patients’ needs, and capture subjects’ contextual changes to help us better understand the role of herbal medicines in the treatment of T2DM patients. This can be crucial in ensuring the best treatment outcomes for T2DM patients while also providing support to biomedical medical providers that Chinese herbal medicine can be effective in supporting these patients. With new cases of T2DM on the rise, and growing numbers of patients with poorly managed disease, further research ought to be conducted to continue to support the claim that Chinese herbal medicine has a role in effectively treating T2DM patients in conjunction with biomedical medical treatment.

## Data Availability

All data produced in the present work are contained in the manuscript.

---

**Supplemental table.**
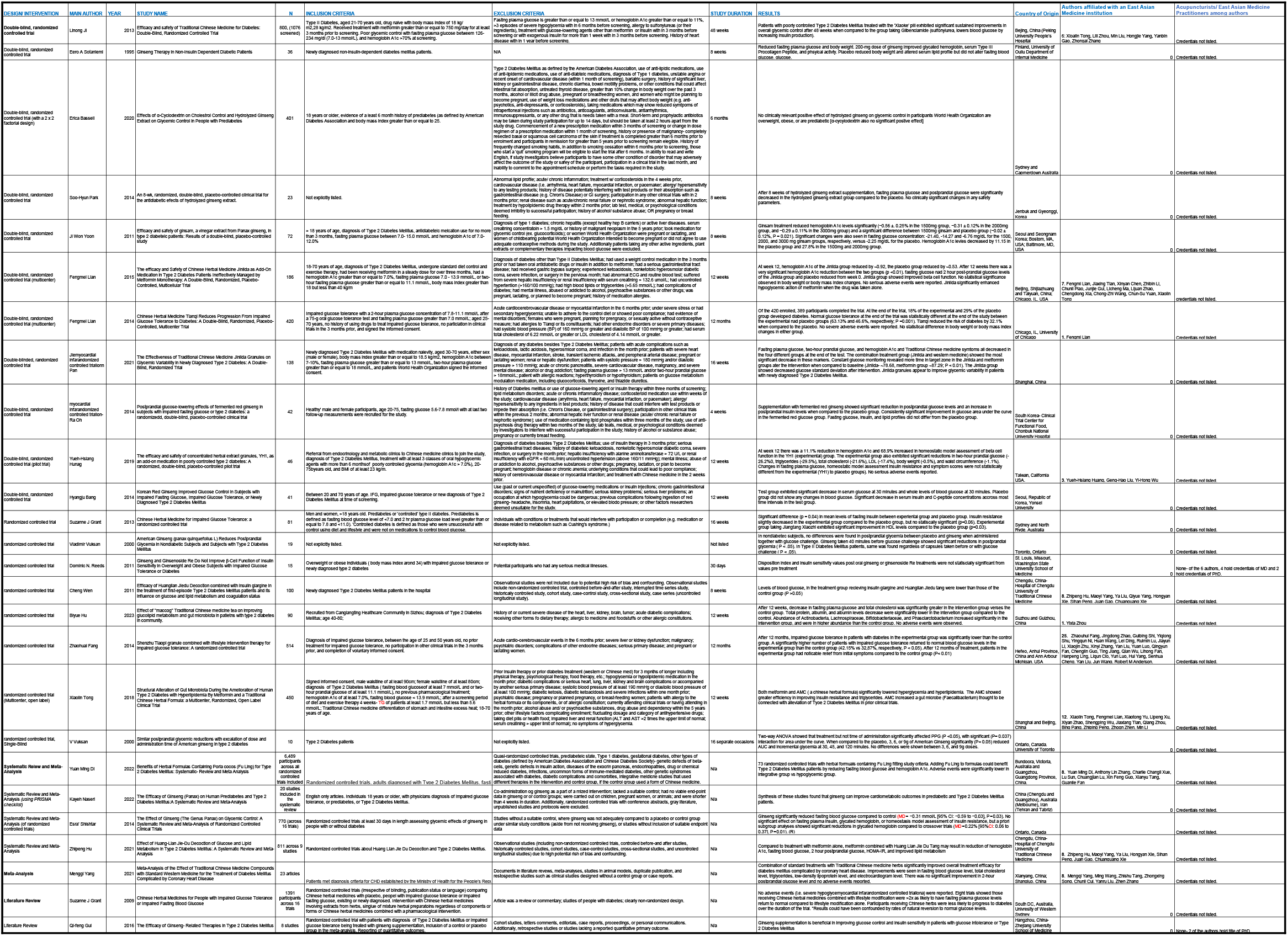
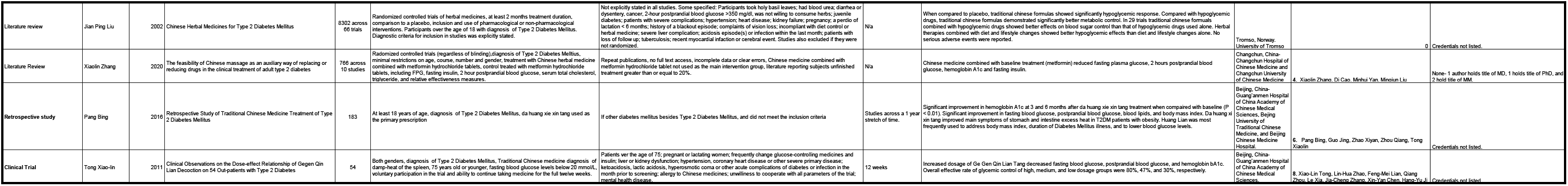

